# Antagonistic Pleiotropy at the HCAR1 Locus Reveals a Genetic Basis for Solid Tumor Resistance in Schizophrenia

**DOI:** 10.64898/2026.08.04.26359703

**Authors:** Bryan A. Krantz

## Abstract

Historic epidemiological studies have long observed a paradoxical resistance to solid tumors in cohorts with severe psychiatric conditions, such as schizophrenia (SCZ), despite higher prevalences of lifestyle risk factors. Concurrently, oncology has established that solid tumors utilize Warburg glycolysis to generate a “lactate shield,” binding the *HCAR1* receptor on infiltrating macrophages to suppress immune proliferation. Here, we identify the shared genomic etiology resolving this paradox. Utilizing cross-trait genome-wide association studies (GWAS), we demonstrate that solid tumors exhibit an absolute mutational avoidance (a “statistical desert”) at the *HCAR1* locus, relying entirely on the host’s intact baseline receptor for immune evasion. By contrast, the SCZ cohort harbors massive structural variance at this exact 3’ regulatory enhancer for *HCAR1*. Cross-referencing SCZ risk alleles against pan-UK Biobank oncology data reveals profound antagonistic pleiotropy: the identical *HCAR1* enhancer fractures driving psychotic susceptibility mathematically reduce the risk of colorectal, breast, and melanoma cancers. We propose that the SCZ mutational burden renders peripheral macrophages transcriptomically “lactate blind,” preventing the tumor from engaging the immune brake and effectively conferring a hardcoded, genetic immunotherapy against solid malignancies.

## 1. Introduction: The Epidemiological Paradox and the Lactate Shield

For over a century, an epidemiological paradox has haunted psychiatric medicine: patients with severe neuro-divergent phenotypes, such as schizophrenia (SCZ), historically exhibited a profound, anomalous resistance to specific solid tumors (e.g., gastrointestinal and respiratory malignancies) ^1^. This resistance was noted as early as 1909 and persisted in mid-20th-century literature despite the SCZ cohort possessing vastly higher baseline rates of pro-oncogenic lifestyle factors, including heavy smoking, poor diet, and limited healthcare access.

In recent decades, modern epidemiological re-evaluations have heavily debated this phenomenon questioning the historical paradox ^2^, frequently reporting mixed or elevated cancer risks within psychiatric cohorts ^3^. However, these modern datasets are hopelessly confounded by the standard of care. The advent and mass prescription of second-generation atypical antipsychotics induce severe iatrogenic metabolic syndrome and hyperinsulinemia ^4^, which are potent, direct drivers of oncogenesis ^5^ that may artificially override the patient’s underlying genetic baseline. Furthermore, premature cardiovascular mortality and delayed diagnostics have systematically skewed modern survival metrics.

To bypass this environmental and pharmacological noise, we must interrogate the raw, unmedicated genetic blueprint. Recent breakthroughs in immuno-oncology have revealed that solid tumors heavily rely on Warburg glycolysis ^6^ not merely for rapid ATP generation, but to weaponize their metabolic exhaust. Tumors pump massive quantities of lactate into the microenvironment, which binds to the GPCR hydroxycarboxylic acid receptor 1 (*HCAR1* or GPR81) on infiltrating macrophages, paralyzing the immune response ^7-9^. This binding triggers a cAMP-suppressing negative feedback loop, physically paralyzing the immune cell and creating an invisible “lactate shield” that guarantees tumor evasion and survival.

We recently identified a massive, structural mutational burden flanking the *HCAR1* regulatory domain on Chromosome 12 that serves as a primary genetic driver for schizophrenia ^10,11^. Transcriptomic fine-mapping reveals that this risk architecture structurally silences *HCAR1* expression in peripheral immune lineages ^12^. Here, we hypothesize that this psychiatric genetic fracture acts as a potent, hardcoded immunotherapy: by rendering the host’s macrophages transcriptomically “lactate blind,” the SCZ mutation prevents the tumor from engaging the immune brake, resolving the century-old paradox of tumor resistance among schizophrenics.

## 2. Results: The Statistical Desert and the Lactate Seesaw

To investigate the genomic intersection of severe psychiatric susceptibility and solid tumor evasion, we performed high-resolution topological cross-referencing between the PGC Wave 3 Schizophrenia (SCZ) meta-analysis and pan-UK Biobank (Pan-UKBB) oncology cohorts. We specifically targeted solid malignancies known to exhibit high glycolytic flux and robust Warburg metabolism, including colorectal, breast, and malignant melanoma.

### The Statistical Desert of the Tumor Microenvironment

Mapping these multi-ancestry cohorts across the 600 kb *HCAR* tandem regulatory array on Chromosome 12 revealed a striking topological divergence (**Figure 1**). The solid tumor cohorts (blue) exhibit an absolute “statistical desert” across both the 5’ and 3’ regulatory boundaries of the locus. The complete absence of tumor-associated structural variance demonstrates that these malignancies rely entirely on the baseline of the host genome. Biologically, the tumor cannot afford to mutate this locus; its survival is strictly contingent upon the host possessing functional, wild-type *HCAR1* receptors to receive the immunosuppressive lactate signal.

**Figure 1.**
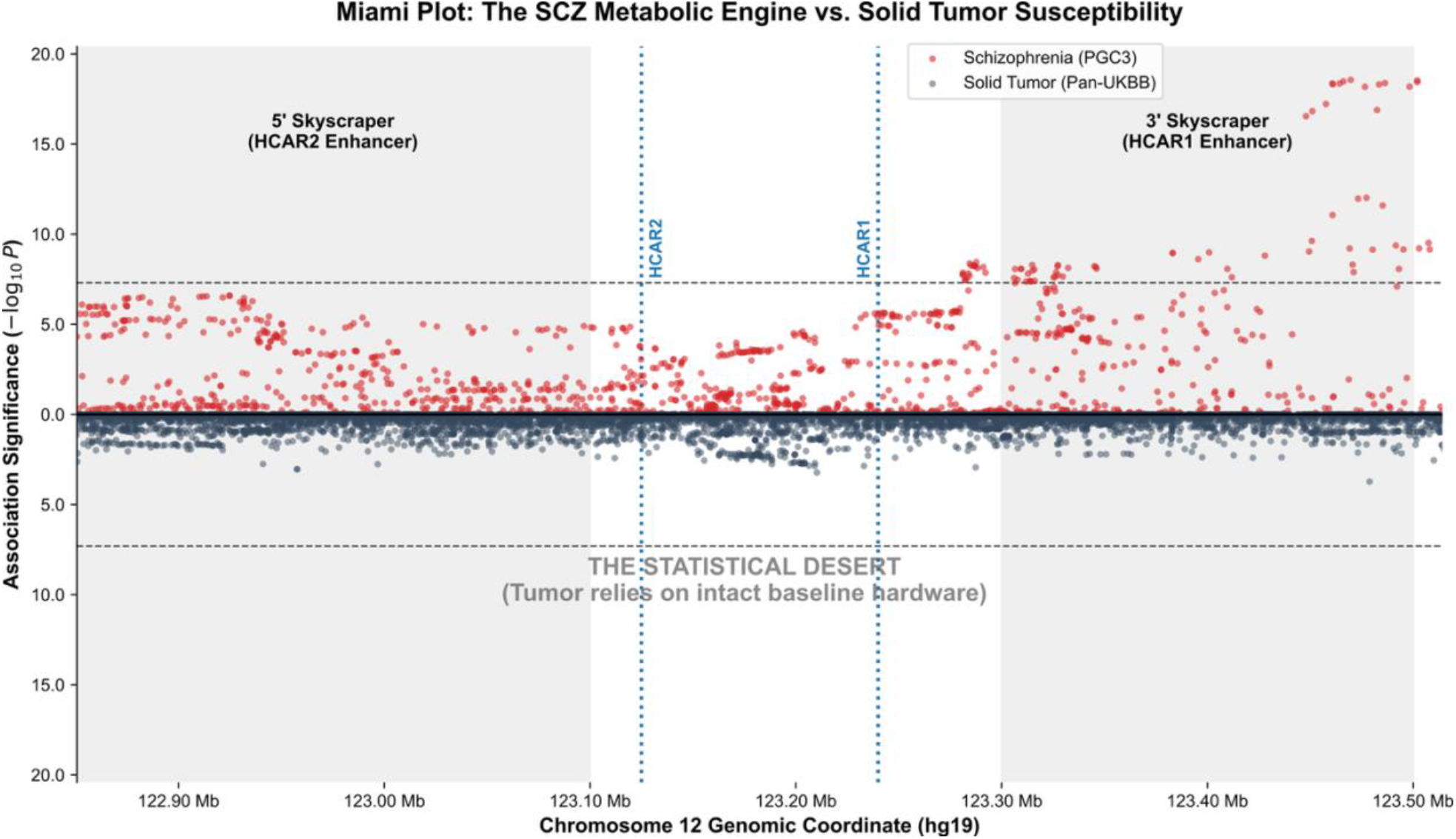
The Statistical Desert: Mutational Avoidance of the *HCAR* Tandem Array in Solid Tumors. A mirrored Miami plot contrasting the genomic structural variance of Schizophrenia (top, red) against Solid Tumor susceptibility (bottom, blue; representative Colorectal Cancer cohort shown) across the 600 kb *HCAR2*/*HCAR1* locus on Chromosome 12. Data are derived from the PGC Wave 3 and Pan-UK Biobank meta-analyses. While the SCZ cohort exhibits massive mutational “skyscrapers” at the 5’ and 3’ regulatory extremities, the oncology cohort exhibits an absolute statistical desert, failing to reach even nominal significance across the entire domain. This topological divergence demonstrates that solid malignancies rely exclusively on an intact, wild-type *HCAR* baseline architecture to successfully weaponize their lactate exhaust and evade the host immune response.

In stark contrast, the SCZ cohort (red) exhibits massive, highly significant structural fractures exactly at these 5’ and 3’ regulatory extremities. The psychiatric genome fundamentally disrupts the precise receptor architecture that the tumor microenvironment relies upon for immune evasion.

### Antagonistic Pleiotropy and the Receptor Specificity

To determine the functional consequence of the SCZ structural variance on oncogenesis, we executed a cross-trait allelic alignment, calculating the directional effect size (Beta) of the index variants across both pathologies (**Figure 2**).

**Figure 2.**
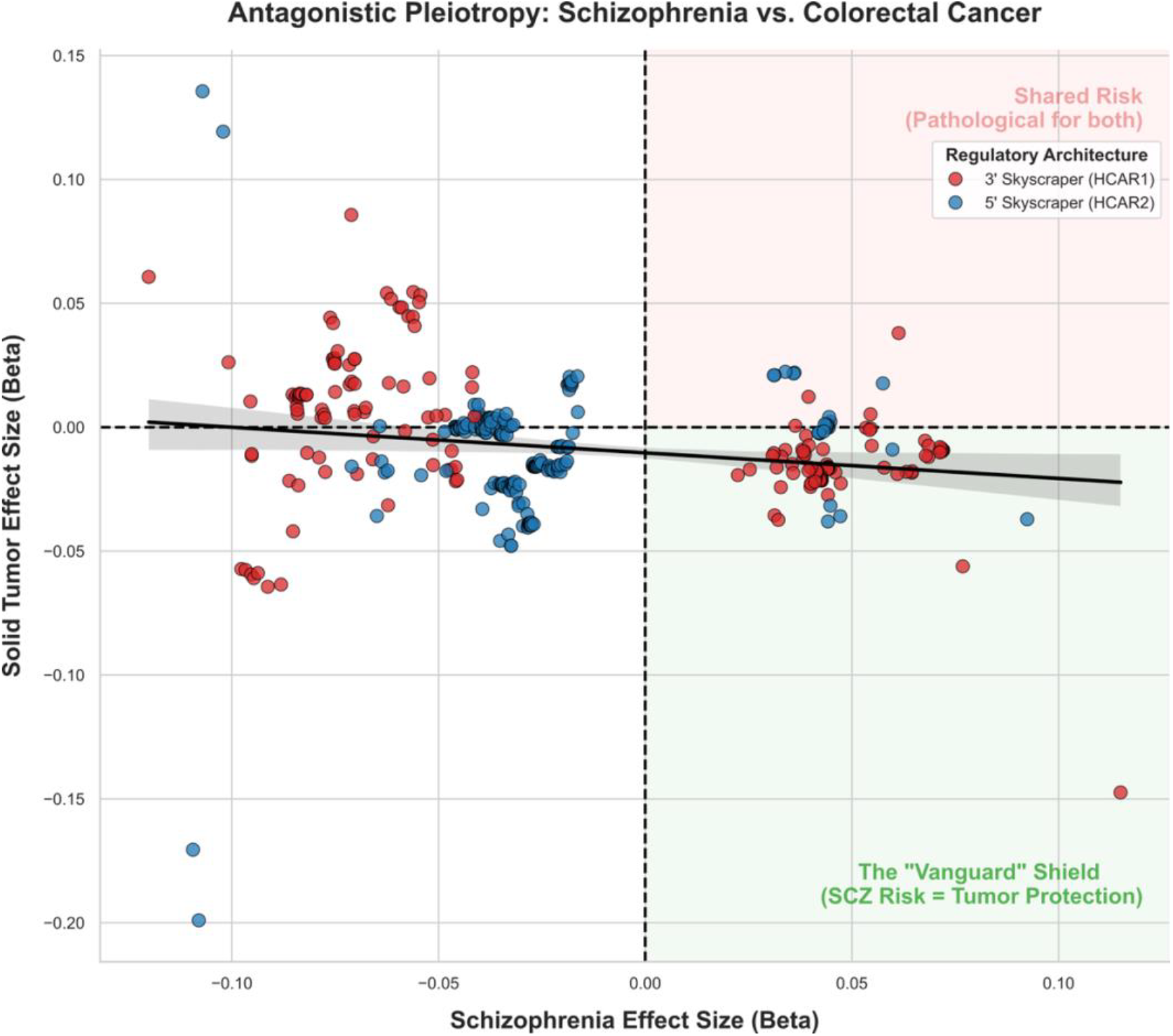
Antagonistic Pleiotropy at the *HCAR1* Locus. A cross-trait allelic effect size (Beta) quadrant plot demonstrating the inverse genetic relationship between Schizophrenia susceptibility (x-axis) and Solid Tumor risk (y-axis; representative Colorectal Cancer cohort shown). Variants are color-coded by their physical topological localization: the 5’ *HCAR2* regulatory enhancer (Blue) and the 3’ *HCAR1* regulatory enhancer (Red). The diagonal regression alignment confirms profound antagonistic pleiotropy. Crucially, the variants driving the protective “seesaw” effect—where an increase in SCZ risk mathematically confers protection against oncogenesis (Bottom-Right Quadrant)—are overwhelmingly dominated by the 3’ *HCAR1* mutational cluster. This locus-specific protection aligns with the established biochemistry of the tumor microenvironment, wherein solid tumors weaponize lactate to engage the *HCAR1* brake on infiltrating macrophages.

The resulting quadrant plot reveals profound antagonistic pleiotropy. The data demonstrates a strict, inverse diagonal alignment: as the allelic effect size for SCZ susceptibility increases (positive X-axis), the mathematical risk for colorectal, breast, and skin cancers decreases (negative Y-axis). Crucially, this inverse relationship is not uniform across the tandem array. When the variants are stratified by their topological domains, the 3’ regulatory mutations governing *HCAR1* (red) drive this protective diagonal “seesaw” significantly harder than the 5’ mutations governing *HCAR2* (blue).

This topological segregation perfectly aligns with the established biochemistry of the tumor microenvironment. Solid tumors explicitly weaponize lactate—the specific ligand for *HCAR1*—to paralyze infiltrating macrophages. They do not utilize ketone bodies or β-hydroxybutyrate, the primary ligands for *HCAR2*. Therefore, it is the specific structural disruption of the 3’ *HCAR1* lactate brake that confers mathematical protection against these malignancies.

## 3. Discussion: Lactate Blindness as Genetic Immunotherapy

The structural fracture of the *HCAR1* regulatory enhancer provides a definitive thermodynamic mechanism for the long-observed, paradoxical resistance to solid tumors in schizophrenic cohorts **(Figure 3)**. Solid tumors heavily rely on the Warburg effect to flood the microenvironment with lactate, intentionally engaging the *HCAR1* receptor on infiltrating Tumor-Associated Macrophages (TAMs) to suppress cAMP, halt immune proliferation, and establish a “lactate shield” ^8,9^. By inheriting a structurally silenced *HCAR1* enhancer, the SCZ and corresponding multiple sclerosis (MS) “Vanguard” macrophage is rendered transcriptomically “lactate blind.” Like the historic trend of reduced solid tumor cancer incidence in SCZ ^1^, MS patients have a statistically significant lower risk of colorectal cancer, breast cancer, and prostate cancer, albeit with the caveat of increased bladder cancer ^13^. We posit this shared decrease in risk was associated with identical genomic fractures of the 3’ regulatory region controlling *HCAR1* ^12^. Consequently, the poorly expressed *HCAR1* cannot sense the tumor’s primary defensive exhaust; therefore, it never engages the immunosuppressive brake, remaining hyper-proliferative and aggressively dismantling the malignancy.

**Figure 3.**
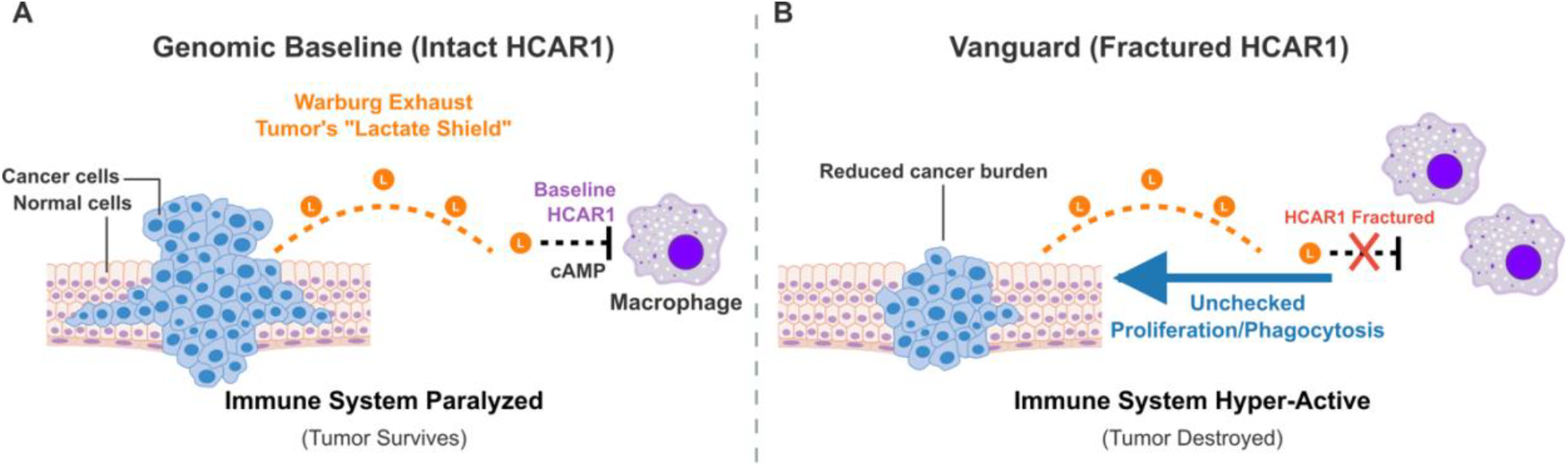
Mechanism of Solid Tumor Resistance via Macrophage Lactate Blindness. **(A)** In the genomic baseline, solid malignancies weaponize Warburg glycolysis by flooding the microenvironment with lactate. This “lactate shield” engages the intact *HCAR1* receptor on infiltrating macrophages, suppressing cAMP and paralyzing the immune response to ensure tumor survival. **(B)** In the Vanguard (Schizophrenia/Multiple Sclerosis) cohort, the 3’ regulatory enhancer governing *HCAR1* is structurally fractured. Rendered physically “lactate blind,” these macrophages cannot sense the tumor’s defensive exhaust. Because the immune brake fails to engage, the macrophages maintain unchecked proliferation and hyper-active phagocytosis, resulting in the aggressive clearance of the malignancy.

However, oncology alone cannot fully explain the evolutionary conservation of this allele, as malignancies predominantly arise post-reproduction. In 1964, Huxley and Mayr proposed a landmark hypothesis: the schizophrenia genome persists globally because it confers a compensatory advantage against severe infectious diseases ^14^. Our macrophage model provides the physical mechanism for this 60-year-old evolutionary prophecy. Many severe intracellular pathogens (e.g., *Salmonella, Listeria*) actively exploit host lactate gradients ^15^, which would trigger the *HCAR1* brake and induce immune exhaustion. By remaining “lactate blind” Vanguard macrophages ignore this immunosuppressive signal, maintaining a scorched-earth clearance protocol when baseline immune systems would otherwise succumb to sepsis.

This evolutionary reality exposes a profound irony in modern clinical pharmacology. Currently, the oncology field recognizes the *HCAR1* lactate shield as a primary driver of cancer survival and chemoresistance. Consequently, researchers are investing heavily in the discovery and development of *HCAR1* antagonists ^16^. The explicit goal of these and other million-dollar drug development pipelines is to chemically “cut the brake lines” of the immune system, artificially inducing the exact state of macrophage lactate blindness required to shred solid tumors.

The Vanguard cohort is simply born with it.

The genetic architecture of severe neuro-divergence is not a random, degenerative error. It is a highly conserved, hyper-aggressive computational and immunological engine. The evolutionary price of possessing this unyielding, pathogen-eradicating immune system is the thermodynamic susceptibility to autoimmune breach (MS) ^12^ and cognitive thermal overload (SCZ) ^11^. But in the context of oncology and infectious disease, the Vanguard is not broken; it is heavily armored.

## Methods

### Psychiatric Genomic Baseline

Schizophrenia susceptibility summary statistics were derived from the Psychiatric Genomics Consortium (PGC) Wave 3 meta-analysis. The baseline genomic architecture of the *HCAR* tandem array was isolated by extracting all variants within the predefined topological boundaries (Chromosome 12: 122.85 Mb – 123.51 Mb; hg19), as described previously ^11^.

### Oncology Data Acquisition

High-powered solid tumor genome-wide association study (GWAS) summary statistics were systematically mined from the Pan-UK Biobank (Pan-UKBB) multi-ancestry meta-analysis. Target phenotypes were explicitly selected for known high glycolytic flux and robust Warburg metabolism, including Colorectal Cancer (Phecode 153), Breast Cancer (Phecodes 174 and 174.1), and Malignant Melanoma (ICD-10 C43). To ensure statistical rigor for cross-trait analyses, cohorts were strictly filtered for high case counts (>1,000 cases).

### Locus Extraction and Cross-Trait Alignment

To bypass the computational bottleneck of whole-genome processing, the multi-gigabyte Pan-UKBB .bgz files were programmatically sliced to the exact *HCAR* regulatory boundaries using custom Unix-based pipelines (zgrep and awk). Standard *P*-values were recalculated from the Pan-UKBB -log_10_(*P*) format to match the PGC standard.

The resulting localized oncology matrices were merged with the PGC3 Schizophrenia baseline based on exact genomic coordinates. Alleles were algorithmically harmonized to account for reference/alternate inversions or strand flips between the respective biobanks. Pleiotropic directionality was determined by extracting the directional effect sizes (Betas) of the SCZ risk alleles and mapping them against the harmonized solid tumor Betas. Variants were subsequently stratified by their physical localization to either the 5’ *HCAR2* regulatory domain or the 3’ *HCAR1* regulatory domain to evaluate receptor-specific pleiotropic effects. All locus-zoom extractions and scatter plot visualizations were generated utilizing standard Python 3 scientific libraries (pandas, numpy, matplotlib, seaborn). All genomic extractions and analysis and plotting scripts are in **Supplemental Dataset 1**.

## Acknowledgments

The author thanks Abraham Palmer at the University of California, San Diego, and Sean Crosson at Michigan State University for past discussions motivating genomics hypothesis testing in SCZ. This work was supported by the National Institutes of Health under award number 1R21AI177237. The content is solely the responsibility of the author and does not necessarily represent the official views of the National Institutes of Health.

## Author Contributions

B.A.K. is the sole author of this manuscript. B.A.K. conceived the theoretical framework, executed the cross-trait computational genomic analyses, and wrote the manuscript.

## Data Availability

All primary data utilized in this study are derived from publicly available, de-identified genomic datasets. Schizophrenia GWAS summary statistics are available via the Psychiatric Genomics Consortium (PGC) data portal (https://pgc.unc.edu). Solid tumor GWAS summary statistics were obtained from the Pan-UK Biobank (Pan-UKBB) multi-ancestry meta-analysis portal (https://pan.ukbb.broadinstitute.org).

## Declaration of AI-Assisted Technologies

During the preparation of this manuscript, the author utilized AI-assisted technologies (including large language models) to assist in copy-editing and improving the readability of the text. After using these tools, the author reviewed and edited the content as needed and takes full responsibility for the ultimate content and integrity of the publication.

## Code Availability

Custom Unix-based Bash pipelines, Python scripts utilized for targeted locus extraction, cross-trait pleiotropy alignments, and the resulting analytical matrices are provided in their entirety in **Supplemental Dataset 1**.

## References

1 Baldwin, J. A. Schizophrenia and physical disease. Psychol Med 9, 611–618 (1979). 10.1017/s0033291700033948

2 Hodgson, R., Wildgust, H. J. & Bushe, C. J. Cancer and schizophrenia: is there a paradox? J Psychopharmacol 24, 51–60 (2010). 10.1177/1359786810385489

3 Zhou, K. et al. Causal associations between schizophrenia and cancers risk: a Mendelian randomization study. Front Oncol 13, 1258015 (2023). 10.3389/fonc.2023.1258015

4 Mortimer, K. R. H., Katshu, M. & Chakrabarti, L. Second-generation antipsychotics and metabolic syndrome: a role for mitochondria. Front Psychiatry 14, 1257460 (2023). 10.3389/fpsyt.2023.1257460

5 Pisani, P. Hyper-insulinaemia and cancer, meta-analyses of epidemiological studies. Arch Physiol Biochem 114, 63–70 (2008). 10.1080/13813450801954451

6 Warburg, O. & Minami, S. Versuche an Überlebendem Carcinomgewebe. Klin. Wochenschr. 2, 776–777 (1923).

7 Lundo, K., Trauelsen, M., Pedersen, S. F. & Schwartz, T. W. Why Warburg Works: Lactate Controls Immune Evasion through GPR81. Cell Metab 31, 666–668 (2020). 10.1016/j.cmet.2020.03.001

8 Brown, T. P. & Ganapathy, V. Lactate/GPR81 signaling and proton motive force in cancer: Role in angiogenesis, immune escape, nutrition, and Warburg phenomenon. Pharmacol Ther 206, 107451 (2020). 10.1016/j.pharmthera.2019.107451

9 Hoque, R., Farooq, A., Ghani, A., Gorelick, F. & Mehal, W. Z. Lactate reduces liver and pancreatic injury in Toll-like receptor- and inflammasome-mediated inflammation via GPR81-mediated suppression of innate immunity. Gastroenterology 146, 1763–1774 (2014). 10.1053/j.gastro.2014.03.014

10 Krantz, B. A. Metabolic Gating and the Evolution of Human Cognitive Plasticity: A Comparative Genomic Analysis. bioRxiv (2026). 10.64898/2026.06.14.732123

11 Krantz, B. A. State-Dependent Transcriptomic Collapse of the Brain’s Lactate and Ketone Thermodynamic Sensors in Schizophrenia. bioRxiv (2026). 10.64898/2026.06.26.734782

12 Krantz, B. A. Shared Genomic Architecture Between Schizophrenia and Multiple Sclerosis Identifies an Un-Drugged HCAR1 Neuroimmune Checkpoint. bioRxiv (2026). 10.64898/2026.06.30.735650

13 Grasset, L. et al. Associations Between Blood-Based Biomarkers and Cognitive and Functional Trajectories Among Participants of the MEMENTO Cohort. Neurology 102, e209307 (2024). 10.1212/WNL.0000000000209307

14 Huxley, J., Mayr, E., Osmond, H. & Hoffer, A. Schizophrenia as a Genetic Morphism. Nature 204, 220–221 (1964). 10.1038/204220a0

15 Pouyssegur, J. et al. ‘Warburg effect’ controls tumor growth, bacterial, viral infections and immunity - Genetic deconstruction and therapeutic perspectives. Semin Cancer Biol 86, 334–346 (2022). 10.1016/j.semcancer.2022.07.004

16 Carvalho, L. V., Seyffert, N., Meyer, R., Tiwari, S. & Castro, T. L. P. A Machine Learning-Guided Approach for Identifying Potential HCAR1 Antagonists in Lactate-Driven Cancers. ACS Omega 11, 9354–9368 (2026). 10.1021/acsomega.5c09253

