## Supplemental Dataset 1 for "Antagonistic Pleiotropy at the HCAR1 Locus Reveals a Genetic Basis for Solid Tumor Resistance in Schizophrenia": cancer_targets.py

```
import pandas as pd
import os

def mine_pan_ukbb_cancer_manifest():
    print("--- Hunting for Solid Tumor GWAS Targets in Pan-UKBB ---")
    
    manifest_file = "Pan-UK Biobank phenotype manifest - phenotype_manifest.csv"
    
    if not os.path.exists(manifest_file):
        print(f"⚠️ Error: Could not find '{manifest_file}'. Ensure it is in the directory.")
        return

    # 1. Load the manifest
    try:
        df = pd.read_csv(manifest_file, low_memory=False)
        print(f"Loaded massive manifest with {len(df)} phenotypes.")
    except Exception as e:
        print(f"⚠️ Error reading manifest: {e}")
        return

    # Ensure column names are lowercase string for easier targeting
    df.columns = [str(col).lower().strip() for col in df.columns]
    
    # 2. Define our Oncology Targets (Solid tumors with high Warburg/glycolytic activity)
    # We want to avoid blood cancers (leukemias) because we specifically want to test 
    # the solid tumor microenvironment (TME) lactate shield.
    target_keywords = [
        'breast cancer', 
        'lung cancer', 
        'colorectal', 
        'malignant melanoma', 
        'prostate cancer',
        'pancreatic'
    ]
    
    # Identify the correct description column
    desc_col = 'description' if 'description' in df.columns else 'trait_name' if 'trait_name' in df.columns else None
    pheno_col = 'phenocode' if 'phenocode' in df.columns else None
    
    if not desc_col or not pheno_col:
        print("⚠️ Warning: Could not identify standard 'description' or 'phenocode' columns.")
        return

    # 3. Apply the Filters
    df[desc_col] = df[desc_col].fillna('')
    df[pheno_col] = df[pheno_col].fillna('')

    # Filter by Keyword (case-insensitive)
    keyword_mask = df[desc_col].str.contains('|'.join(target_keywords), case=False)
    target_df = df[keyword_mask].copy()

    # 4. Filter for Power 
    # Cancer phenotypes can be rare. We need high N to detect the pleiotropy reliably.
    if 'n_cases_white_british' in df.columns:
        target_df = target_df[target_df['n_cases_white_british'] > 1000]
    elif 'n_cases_eur' in df.columns:
        target_df = target_df[target_df['n_cases_eur'] > 1000]

    # 5. Format the Output
    output_cols = [pheno_col, desc_col]
    for col in ['n_cases_eur', 'n_cases_white_british', 'wget_command', 'aws_link']:
        if col in df.columns:
            output_cols.append(col)
            
    final_hits = target_df[output_cols].sort_values(by=output_cols[2], ascending=False) # Sort by case count
    
    # Export the hit-list
    output_csv = "Oncology_Target_GWAS_Files.csv"
    final_hits.to_csv(output_csv, index=False)
    
    print(f"\n✅ Success! Narrowed down to {len(final_hits)} high-powered oncology targets.")
    print(f"📁 Target manifest saved to: {output_csv}")
    print("\n--- PREVIEW OF TOP HITS (Sorted by Case Count) ---")
    print(final_hits[[pheno_col, desc_col]].head(10).to_string(index=False))

if __name__ == "__main__":
    mine_pan_ukbb_cancer_manifest()
```
