## Supplemental Dataset 1 for "Antagonistic Pleiotropy at the HCAR1 Locus Reveals a Genetic Basis for Solid Tumor Resistance in Schizophrenia": cancer_vs_scz_cross_reference.py

```
import pandas as pd
import numpy as np
import glob
import os

# --- CONFIGURE ---
SCZ_FILE = "Tandem_HCAR2_HCAR1_wide_regulatory.tsv"
INPUT_DIR = "./" # Directory where your UKBB_HCAR_Locus_Extracted_*.tsv files are
REGION_5_PRIME_START = 122_850_000
REGION_5_PRIME_END   = 123_100_000
REGION_3_PRIME_START = 123_300_000
REGION_3_PRIME_END   = 123_500_000

def compare_scz_to_cancer():
    print("--- Initiating Oncology vs. SCZ Cross-Reference (Batch Mode) ---")
    
    # 1. Load SCZ Baseline
    if not os.path.exists(SCZ_FILE):
        print(f"⚠️ Error: SCZ baseline '{SCZ_FILE}' not found.")
        return
        
    df_scz = pd.read_csv(SCZ_FILE, sep='\t')
    df_scz['POS'] = pd.to_numeric(df_scz['POS'], errors='coerce')
    
    # 2. Get list of all extracted cancer files
    cancer_files = glob.glob(os.path.join(INPUT_DIR, "UKBB_HCAR_Locus_Extracted_*.tsv"))
    
    if not cancer_files:
        print("⚠️ No extracted cancer files found. Check your directory.")
        return

    for cancer_file in cancer_files:
        filename = os.path.basename(cancer_file)
        print(f"\nProcessing: {filename}")
        
        df_cancer = pd.read_csv(cancer_file, sep='\t')
        df_cancer['POS'] = pd.to_numeric(df_cancer['POS'], errors='coerce')
        
        # Merge
        merged = pd.merge(df_scz, df_cancer, on='POS', suffixes=('_SCZ', '_CANCER'), how='inner')
        
        if merged.empty:
            print("  ⚠️ No overlap found for this phenotype.")
            continue
            
        # 3. Analyze Allele Alignment and Effect
        results = []
        for _, row in merged.iterrows():
            # Align Alleles (Pandas appended _CANCER to the UKBB columns during the merge)
            scz_a1 = str(row['A1_SCZ']).upper()
            scz_a2 = str(row['A2_SCZ']).upper()
            c_a1 = str(row['A1_CANCER']).upper()
            c_a2 = str(row['A2_CANCER']).upper()
            
            scz_beta = float(row['BETA_SCZ'])
            c_beta = float(row['BETA_CANCER'])
            
            # Flip Beta if alleles are reversed
            if scz_a1 == c_a1 and scz_a2 == c_a2:
                aligned_beta = c_beta
            elif scz_a1 == c_a2 and scz_a2 == c_a1:
                aligned_beta = -c_beta
            else:
                continue # Skip ambiguous/strand-flipped
                
            # Determine relationship
            # If SCZ Risk (Beta > 0) has Negative Cancer Beta, it's Protective
            if scz_beta > 0 and aligned_beta < 0:
                rel = "PROTECTIVE (SCZ Risk reduces Cancer Risk)"
            elif scz_beta < 0 and aligned_beta > 0:
                rel = "PROTECTIVE (SCZ Protective reduces Cancer Risk)"
            elif (scz_beta > 0 and aligned_beta > 0) or (scz_beta < 0 and aligned_beta < 0):
                rel = "SHARED RISK (Same Direction)"
            else:
                rel = "Neutral"
            
            # Determine Region
            pos = row['POS']
            if REGION_5_PRIME_START <= pos <= REGION_5_PRIME_END:
                region = "5' Skyscraper (HCAR2)"
            elif REGION_3_PRIME_START <= pos <= REGION_3_PRIME_END:
                region = "3' Skyscraper (HCAR1)"
            else:
                region = "Intergenic"
            
            results.append({
                'rsID': row['ID_SCZ'], 'POS': pos, 'Region': region,
                'PVAL_SCZ': row['PVAL_SCZ'], 'SCZ_BETA': scz_beta,
                'PVAL_CANCER': row['PVAL_CANCER'], 'CANCER_BETA': aligned_beta,
                'Effect': rel
            })
            
        # 4. Save results per phenotype
        if results:
            res_df = pd.DataFrame(results)
            # Sort by SCZ significance so the most important Vanguard hits are at the top
            res_df = res_df.sort_values(by='PVAL_SCZ', ascending=True)
            
            out_name = f"Cross_Ref_{filename}"
            res_df.to_csv(out_name, sep='\t', index=False)
            print(f"  ✅ Exported results: {out_name}")
            
            protective_count = len(res_df[res_df['Effect'].str.contains('PROTECTIVE')])
            total_count = len(res_df)
            print(f"  Found {protective_count} protective hits out of {total_count} perfectly aligned variants.")

if __name__ == "__main__":
    compare_scz_to_cancer()
```
