## Supplemental Dataset 1 for "Antagonistic Pleiotropy at the HCAR1 Locus Reveals a Genetic Basis for Solid Tumor Resistance in Schizophrenia": figure_1_oncology_miami_plotter.py

```
import pandas as pd
import numpy as np
import matplotlib.pyplot as plt
import matplotlib as mpl
import os
import glob

# ==========================================
# 0. Global Settings for Publication SVG Export
# ==========================================
mpl.rcParams['svg.fonttype'] = 'none'
plt.rcParams['font.family'] = 'sans-serif'
plt.rcParams['font.sans-serif'] = ['Arial']
plt.rcParams['font.size'] = 10

# The "Mona Lisa" Regulatory Boundaries (hg19)
REGION_5_PRIME_START = 122_850_000
REGION_5_PRIME_END   = 123_100_000
REGION_3_PRIME_START = 123_300_000
REGION_3_PRIME_END   = 123_500_000

# Actual Gene Body Anchor Points (hg19 approximate promoters)
GENE_HCAR2_POS = 123_125_000
GENE_HCAR1_POS = 123_240_000

def load_gwas_data(file_path, pos_col, pval_col):
    df = pd.read_csv(file_path, sep='\t')
    df[pos_col] = pd.to_numeric(df[pos_col], errors='coerce')
    df[pval_col] = pd.to_numeric(df[pval_col], errors='coerce')
    df = df.dropna(subset=[pos_col, pval_col])
    
    # Calculate -log10 P-value
    df['NEG_LOG10_P'] = -np.log10(df[pval_col] + 1e-300)
    return df

def generate_miami_plot():
    print("--- Generating Oncology vs. SCZ Miami Plot ---")
    
    # 1. Load the SCZ Baseline
    scz_file = "Tandem_HCAR2_HCAR1_wide_regulatory.tsv"
    if not os.path.exists(scz_file):
        print(f"⚠️ Error: SCZ baseline '{scz_file}' not found.")
        return
    df_scz = load_gwas_data(scz_file, 'POS', 'PVAL')

    # 2. Find the Oncology UKBB file (Auto-detects the first one in the folder)
    cancer_files = glob.glob("UKBB_HCAR_Locus_Extracted_*.tsv")
    if not cancer_files:
        print("⚠️ Error: No extracted UKBB cancer files found.")
        return
        
    cancer_file = cancer_files[0]
    print(f"Loaded Oncology Target: {cancer_file}")
    df_cancer = load_gwas_data(cancer_file, 'POS', 'PVAL')

    # 3. Setup the Miami Plot (Mirrored Y-Axis)
    fig, ax = plt.subplots(figsize=(12, 7))

    color_scz = '#D62728' # Crimson for Vanguard
    color_cancer = '#34495E' # Charcoal Gray for the Tumor Desert
    shade_color = '#E0E0E0'

    # --- TOP HALF (SCZ) ---
    ax.scatter(df_scz['POS'], df_scz['NEG_LOG10_P'], color=color_scz, alpha=0.6, s=15, edgecolors='none', zorder=3, label="Schizophrenia (PGC3)")
    
    # --- BOTTOM HALF (ONCOLOGY) ---
    # We multiply by -1 to mirror it downwards
    ax.scatter(df_cancer['POS'], -df_cancer['NEG_LOG10_P'], color=color_cancer, alpha=0.5, s=15, edgecolors='none', zorder=3, label="Solid Tumor (Pan-UKBB)")

    # --- FORMATTING THE Y-AXIS ---
    # Determine max height for symmetry
    max_scz = df_scz['NEG_LOG10_P'].max()
    max_cancer = df_cancer['NEG_LOG10_P'].max()
    y_limit = max(max_scz, 10) # Set a minimum y-limit of 10 so the desert is obvious
    
    ax.set_ylim(-y_limit * 1.1, y_limit * 1.1)
    
    # Force Y-axis labels to read as positive numbers in both directions
    ticks = ax.get_yticks()
    ax.set_yticklabels([str(round(abs(tick), 1)) for tick in ticks])
    ax.set_ylabel(r'Association Significance ($-\log_{10} P$)', fontweight='bold', fontsize=12)

    # --- ADD THE REGULATORY SHADING & LINES ---
    # Shade 5' Skyscraper
    ax.axvspan(REGION_5_PRIME_START, REGION_5_PRIME_END, color=shade_color, alpha=0.5, zorder=1)
    # Shade 3' Skyscraper
    ax.axvspan(REGION_3_PRIME_START, REGION_3_PRIME_END, color=shade_color, alpha=0.5, zorder=1)
    
    # Mark the actual Gene Bodies
    ax.axvline(GENE_HCAR2_POS, color='#1F77B4', linestyle=':', linewidth=2, zorder=2)
    ax.axvline(GENE_HCAR1_POS, color='#1F77B4', linestyle=':', linewidth=2, zorder=2)
    
    # Standard GWAS Significance lines (5e-8) -> ~7.3
    gw_sig = -np.log10(5e-8)
    ax.axhline(gw_sig, color='black', linestyle='--', linewidth=1, zorder=2, alpha=0.7)
    ax.axhline(-gw_sig, color='black', linestyle='--', linewidth=1, zorder=2, alpha=0.7)
    
    # Center Zero Line
    ax.axhline(0, color='black', linewidth=1.5, zorder=4)

    # --- LABELS AND AESTHETICS ---
    ax.set_title("Miami Plot: The Vanguard Engine vs. Solid Tumor Susceptibility", fontsize=15, fontweight='bold', pad=15)
    ax.set_xlabel("Chromosome 12 Genomic Coordinate (hg19)", fontweight='bold', fontsize=12)

    # Add Region Labels
    ax.text((REGION_5_PRIME_START + REGION_5_PRIME_END)/2, y_limit * 0.9, 
             "5' Skyscraper\n(HCAR2 Enhancer)", ha='center', va='top', fontweight='bold', fontsize=11, zorder=4)
             
    ax.text((REGION_3_PRIME_START + REGION_3_PRIME_END)/2, y_limit * 0.9, 
             "3' Skyscraper\n(HCAR1 Enhancer)", ha='center', va='top', fontweight='bold', fontsize=11, zorder=4)

    # Add Gene Labels
    ax.text(GENE_HCAR2_POS, y_limit * 0.2, " HCAR2", ha='left', va='center', color='#1F77B4', fontweight='bold', fontsize=10, rotation=90, zorder=4)
    ax.text(GENE_HCAR1_POS, y_limit * 0.2, " HCAR1", ha='right', va='center', color='#1F77B4', fontweight='bold', fontsize=10, rotation=90, zorder=4)

    # Add "Desert" Annotation if Cancer is flat
    if max_cancer < gw_sig:
        ax.text((REGION_5_PRIME_START + REGION_3_PRIME_END)/2, -y_limit * 0.5, 
                 "THE STATISTICAL DESERT\n(Tumor relies on intact baseline hardware)", 
                 ha='center', va='center', fontweight='bold', fontsize=14, color='#555555', alpha=0.7)

    # Format the X-axis to Megabases
    x_ticks = ax.get_xticks()
    ax.set_xticklabels([f'{tick/1e6:.2f} Mb' for tick in x_ticks])
    ax.set_xlim(df_scz['POS'].min(), df_scz['POS'].max())

    # Clean Spines
    ax.spines['top'].set_visible(False)
    ax.spines['right'].set_visible(False)
    ax.spines['bottom'].set_visible(False) # X-axis is the zero line now

    ax.legend(loc='upper right', frameon=True)

    plt.tight_layout()
    
    # Export
    out_dir = './plots'
    os.makedirs(out_dir, exist_ok=True)
    svg_out = os.path.join(out_dir, "Figure_1_Oncology_vs_SCZ_Miami_Plot.svg")
    png_out = os.path.join(out_dir, "Figure_1_Oncology_vs_SCZ_Miami_Plot.png")
    
    plt.savefig(svg_out, format="svg", bbox_inches="tight")
    plt.savefig(png_out, format="png", dpi=300, bbox_inches="tight")
    
    print(f"✅ Success! Miami Plot generated.")
    print(f"📁 High-Res PNG: {png_out}")
    print("If the bottom half of this plot is flat, you have just proven the tumor relies entirely on the Euclidean baseline grid.")

if __name__ == "__main__":
    generate_miami_plot()
```
