## Supplemental Dataset 1 for "Antagonistic Pleiotropy at the HCAR1 Locus Reveals a Genetic Basis for Solid Tumor Resistance in Schizophrenia": figure_2_antagonistic_quadrant_fixed.py

```
import pandas as pd
import numpy as np
import matplotlib.pyplot as plt
import seaborn as sns
import matplotlib as mpl
import os

# ==========================================
# Global Font Settings for Publication
# ==========================================
mpl.rcParams['svg.fonttype'] = 'none'
plt.rcParams['font.family'] = 'sans-serif'
plt.rcParams['font.sans-serif'] = ['Arial']
plt.rcParams['font.size'] = 11

def generate_pleiotropy_quadrant_plot():
    print("--- Generating Figure 2: Antagonistic Pleiotropy Quadrant Plot ---")
    
    # Using Colorectal Cancer (153) as the prime example due to highest hit count
    input_file = "Cross_Ref_UKBB_HCAR_Locus_Extracted_phecode-153-both_sexes_chr12.tsv"
    
    if not os.path.exists(input_file):
        print(f"⚠️ Error: Could not find '{input_file}'.")
        return
        
    df = pd.read_csv(input_file, sep='\t')
    
    # Filter out Intergenic noise to focus purely on the two regulatory skyscrapers
    df = df[df['Region'].isin(["3' Skyscraper (HCAR1)", "5' Skyscraper (HCAR2)"])]
    
    # Filter for variants that have at least some nominal significance in SCZ 
    # to clean up the center blob of zero-effect SNPs
    df = df[df['PVAL_SCZ'] < 0.05]

    sns.set_theme(style="whitegrid", context="paper")
    fig, ax = plt.subplots(figsize=(9, 8))
    
    # Define exact colors
    color_dict = {
        "3' Skyscraper (HCAR1)": "#D62728", # Crimson Red
        "5' Skyscraper (HCAR2)": "#1F77B4"  # Cobalt Blue
    }

    # 1. Plot the Data First (So we can extract the true axis limits)
    sns.scatterplot(
        data=df,
        x='SCZ_BETA',
        y='CANCER_BETA',
        hue='Region',
        palette=color_dict,
        s=50,
        alpha=0.75,
        edgecolor='black',
        linewidth=0.5,
        zorder=3,
        ax=ax
    )

    # 2. Add Linear Regression Line to show the overall inverse trend
    sns.regplot(
        data=df, x='SCZ_BETA', y='CANCER_BETA', scatter=False, 
        color='black', line_kws={'linewidth': 2, 'zorder': 4}, ax=ax
    )

    # Extract the absolute data limits to draw precise bounding boxes
    x_min, x_max = ax.get_xlim()
    y_min, y_max = ax.get_ylim()

    # 3. Draw Quadrant Backgrounds (Using absolute DATA coordinates)
    # Bottom-Right: The Vanguard Shield (SCZ Risk, Cancer Protective)
    ax.fill_between([0, x_max], 0, y_min, color='green', alpha=0.05, zorder=0)
    ax.text(x_max * 0.95, y_min * 0.9, 
            "The Vanguard Shield\n(SCZ Risk = Tumor Protection)", 
            ha='right', va='bottom', fontsize=12, fontweight='bold', color='#2CA02C', alpha=0.8)

    # Top-Right: Shared Risk (Empty theoretically)
    ax.fill_between([0, x_max], 0, y_max, color='red', alpha=0.05, zorder=0)
    ax.text(x_max * 0.95, y_max * 0.9, 
            "Shared Risk\n(Pathological for both)", 
            ha='right', va='top', fontsize=12, fontweight='bold', color='#D62728', alpha=0.4)

    # 4. Add Center Crosshairs locked to zero
    ax.axhline(0, color='black', linewidth=1.5, linestyle='--', zorder=1)
    ax.axvline(0, color='black', linewidth=1.5, linestyle='--', zorder=1)

    # 5. Formatting
    ax.set_title("Antagonistic Pleiotropy: Schizophrenia vs. Colorectal Cancer", fontsize=15, fontweight='bold', pad=15)
    ax.set_xlabel("Schizophrenia Effect Size (Beta)", fontsize=13, fontweight='bold', labelpad=10)
    ax.set_ylabel("Solid Tumor Effect Size (Beta)", fontsize=13, fontweight='bold', labelpad=10)
    
    # Re-apply the original limits so the shading doesn't distort the graph size
    ax.set_xlim(x_min, x_max)
    ax.set_ylim(y_min, y_max)
    
    # Legend formatting
    legend = ax.legend(title="Regulatory Architecture", loc='upper right', frameon=True, bbox_to_anchor=(1.0, 1.0))
    legend.get_title().set_fontweight('bold')
    
    sns.despine()
    plt.tight_layout()
    
    out_dir = './plots'
    os.makedirs(out_dir, exist_ok=True)
    svg_out = os.path.join(out_dir, "Figure_2_Antagonistic_Quadrant_Fixed.svg")
    png_out = os.path.join(out_dir, "Figure_2_Antagonistic_Quadrant_Fixed.png")
    
    plt.savefig(svg_out, format="svg", bbox_inches="tight")
    plt.savefig(png_out, format="png", dpi=300, bbox_inches="tight")
    
    print(f"✅ Success! Quadrant Plot geometry fixed.")
    print(f"📁 High-Res PNG: {png_out}")

if __name__ == "__main__":
    generate_pleiotropy_quadrant_plot()
```
