## Supplemental Dataset 1 for "Antagonistic Pleiotropy at the HCAR1 Locus Reveals a Genetic Basis for Solid Tumor Resistance in Schizophrenia": figure_3_oncology_schematic.py

```
import os

def generate_oncology_schematic():
    print("--- Generating Figure 3: The Vanguard Macrophage in Oncology ---")
    
    # SVG Configuration (10 inches wide by 5 inches tall)
    width_in = 10
    height_in = 5
    dpi = 100
    width_px = int(width_in * dpi)
    height_px = int(height_in * dpi)
    
    # Color palette
    colors = {
        'macrophage_wt': '#BDC3C7',     # Sleepy Gray
        'macrophage_van': '#1F77B4',    # Active Cobalt Blue
        'tumor': '#D62728',             # Malignant Red
        'lactate': '#FF7F0E',           # Bright Orange
        'text_dark': '#333333',
        'bg_color': '#FDFDFD',
        'line_gray': '#95A5A6'
    }

    svg_content = f"""<?xml version="1.0" encoding="UTF-8" standalone="no"?>
<svg xmlns="http://www.w3.org/2000/svg" viewBox="0 0 {width_px} {height_px}" width="{width_in}in" height="{height_in}in">
    
    <defs>
        <!-- Arrowhead markers -->
        <marker id="arrow-dark" viewBox="0 0 10 10" refX="9" refY="5" markerWidth="6" markerHeight="6" orient="auto-start-reverse">
            <path d="M 0 0 L 10 5 L 0 10 z" fill="{colors['text_dark']}"/>
        </marker>
        <marker id="arrow-blue" viewBox="0 0 10 10" refX="9" refY="5" markerWidth="6" markerHeight="6" orient="auto-start-reverse">
            <path d="M 0 0 L 10 5 L 0 10 z" fill="{colors['macrophage_van']}"/>
        </marker>
        
        <!-- Drop shadow for 3D depth -->
        <filter id="drop-shadow" x="-20%" y="-20%" width="140%" height="140%">
            <feDropShadow dx="2" dy="4" stdDeviation="3" flood-opacity="0.2"/>
        </filter>
        
        <!-- Lactate Molecule Pattern -->
        <g id="lactate">
            <circle cx="0" cy="0" r="6" fill="{colors['lactate']}"/>
            <text x="0" y="2.5" font-family="Arial" font-size="7" font-weight="bold" fill="white" text-anchor="middle">L</text>
        </g>
    </defs>

    <!-- Background -->
    <rect width="100%" height="100%" fill="{colors['bg_color']}"/>

    <!-- Center Divider Line -->
    <line x1="500" y1="50" x2="500" y2="450" stroke="{colors['line_gray']}" stroke-width="2" stroke-dasharray="8,8"/>

    <!-- ========================================== -->
    <!-- PANEL A: EUCLIDEAN MAINTAINER (Tumor Wins) -->
    <!-- ========================================== -->
    <g id="panel-a" transform="translate(30, 0)">
        <text x="220" y="40" font-family="Arial" font-size="16" font-weight="bold" fill="{colors['text_dark']}" text-anchor="middle">A. Euclidean Baseline (Intact HCAR1)</text>
        
        <!-- Solid Tumor -->
        <circle cx="100" cy="250" r="50" fill="{colors['tumor']}" filter="url(#drop-shadow)"/>
        <circle cx="70" cy="220" r="30" fill="{colors['tumor']}" filter="url(#drop-shadow)"/>
        <circle cx="130" cy="280" r="35" fill="{colors['tumor']}" filter="url(#drop-shadow)"/>
        <text x="100" y="255" font-family="Arial" font-size="14" font-weight="bold" fill="white" text-anchor="middle">Solid Tumor</text>
        
        <!-- Warburg Exhaust (Lactate Shield) -->
        <path d="M 160 250 Q 220 200 280 250" stroke="{colors['lactate']}" stroke-width="3" stroke-dasharray="5,5" fill="none"/>
        <use href="#lactate" x="180" y="225"/>
        <use href="#lactate" x="220" y="210"/>
        <use href="#lactate" x="260" y="225"/>
        <text x="220" y="190" font-family="Arial" font-size="12" font-weight="bold" fill="{colors['lactate']}" text-anchor="middle">Warburg Exhaust</text>

        <!-- Wild-Type Macrophage -->
        <path d="M 330 250 C 330 210, 390 210, 390 250 C 390 290, 330 290, 330 250 Z" fill="{colors['macrophage_wt']}" filter="url(#drop-shadow)"/>
        <text x="360" y="255" font-family="Arial" font-size="12" font-weight="bold" fill="{colors['text_dark']}" text-anchor="middle">Macrophage</text>
        
        <!-- Intact HCAR1 Receptor -->
        <path d="M 335 250 L 320 240 L 320 260 Z" fill="#9B59B6"/>
        <use href="#lactate" x="315" y="250"/>
        <text x="310" y="280" font-family="Arial" font-size="10" font-weight="bold" fill="#9B59B6" text-anchor="end">HCAR1 Intact</text>
        
        <!-- Consequence -->
        <text x="360" y="320" font-family="Arial" font-size="14" font-weight="bold" fill="{colors['text_dark']}" text-anchor="middle">Immune System Paralyzed</text>
        <text x="360" y="340" font-family="Arial" font-size="12" fill="{colors['text_dark']}" text-anchor="middle">(Tumor Survives)</text>
    </g>

    <!-- ========================================== -->
    <!-- PANEL B: VANGUARD SCOUT (Tumor Dies)       -->
    <!-- ========================================== -->
    <g id="panel-b" transform="translate(530, 0)">
        <text x="220" y="40" font-family="Arial" font-size="16" font-weight="bold" fill="{colors['text_dark']}" text-anchor="middle">B. Vanguard Scout ("Lactate Blind")</text>
        
        <!-- Solid Tumor (Dying/Shrinking) -->
        <circle cx="100" cy="250" r="40" fill="{colors['tumor']}" opacity="0.6" filter="url(#drop-shadow)"/>
        <circle cx="75" cy="225" r="20" fill="{colors['tumor']}" opacity="0.6" filter="url(#drop-shadow)"/>
        <circle cx="125" cy="275" r="25" fill="{colors['tumor']}" opacity="0.6" filter="url(#drop-shadow)"/>
        <text x="100" y="255" font-family="Arial" font-size="14" font-weight="bold" fill="white" text-anchor="middle">Solid Tumor</text>
        
        <!-- Warburg Exhaust (Lactate Shield Failing) -->
        <path d="M 150 250 Q 210 200 270 250" stroke="{colors['lactate']}" stroke-width="3" stroke-dasharray="5,5" fill="none"/>
        <use href="#lactate" x="170" y="225"/>
        <use href="#lactate" x="210" y="210"/>
        <use href="#lactate" x="250" y="225"/>

        <!-- Vanguard Macrophage (Aggressive/Active) -->
        <path d="M 330 250 C 320 200, 400 200, 390 250 C 410 290, 310 290, 330 250 Z" fill="{colors['macrophage_van']}" filter="url(#drop-shadow)"/>
        <text x="360" y="255" font-family="Arial" font-size="12" font-weight="bold" fill="white" text-anchor="middle">Macrophage</text>
        
        <!-- Fractured HCAR1 Receptor -->
        <line x1="320" y1="240" x2="335" y2="260" stroke="#E74C3C" stroke-width="3"/>
        <line x1="335" y1="240" x2="320" y2="260" stroke="#E74C3C" stroke-width="3"/>
        <text x="310" y="280" font-family="Arial" font-size="10" font-weight="bold" fill="#E74C3C" text-anchor="end">HCAR1 Fractured</text>
        
        <!-- Attack Arrow -->
        <path d="M 320 250 L 160 250" stroke="{colors['macrophage_van']}" stroke-width="5" fill="none" marker-end="url(#arrow-blue)"/>
        <text x="240" y="270" font-family="Arial" font-size="12" font-weight="bold" fill="{colors['macrophage_van']}" text-anchor="middle">Unchecked Phagocytosis</text>

        <!-- Consequence -->
        <text x="360" y="320" font-family="Arial" font-size="14" font-weight="bold" fill="{colors['macrophage_van']}" text-anchor="middle">Immune System Hyper-Active</text>
        <text x="360" y="340" font-family="Arial" font-size="12" fill="{colors['text_dark']}" text-anchor="middle">(Tumor Destroyed)</text>
    </g>

</svg>
"""
    
    filename = "./plots/Figure_3_Oncology_Schematic.svg"
    with open(filename, "w", encoding="utf-8") as f:
        f.write(svg_content)
        
    print(f"✅ Success! Generated Figure 3 schematic: {filename}")
    print("You now have the complete 3-Figure package for the Brief Communication.")

if __name__ == "__main__":
    generate_oncology_schematic()
```
