## Supplemental Dataset 1 for "Antagonistic Pleiotropy at the HCAR1 Locus Reveals a Genetic Basis for Solid Tumor Resistance in Schizophrenia": README.docx

**Oncology vs. Vanguard: Genomic Extraction & Cross-Trait Pleiotropy Pipeline**

**Author:** Bryan A. Krantz

**Overview**

This repository contains the computational pipeline utilized to investigate the genomic intersection of severe psychiatric susceptibility (Schizophrenia) and solid tumor evasion.

To test the hypothesis that the *HCAR1* (GPR81) lactate receptor acts as an un-drugged immune evasion shield for solid tumors, this pipeline extracts the tandem *HCAR2/HCAR1* regulatory locus from the Pan-UK Biobank (Pan-UKBB) multi-ancestry meta-analysis. It cross-references these oncology datasets against the Psychiatric Genomics Consortium (PGC) Wave 3 Schizophrenia baseline to map topological avoidance (The "Statistical Desert") and calculate antagonistic pleiotropy (The "Lactate Seesaw").

**Directory Structure & Pipeline Workflow**

**Phase 1: Phenotype Targeting & Acquisition**

- **Script:** cancer_targets.py
- **Input:** Pan-UK Biobank phenotype manifest - phenotype_manifest.csv
- **Process:** Programmatically mines the massive Pan-UKBB manifest for solid malignancies known to exhibit high glycolytic flux and robust Warburg metabolism (e.g., colorectal, breast, melanoma). Filters for high-powered case counts to ensure statistical rigor.
- **Output:** Oncology_Target_GWAS_Files.csv (Contains the direct wget URLs used to acquire the multi-gigabyte .tsv.bgz summary statistics files).

**Phase 2: High-Resolution Genomic Slicing**

- **Process:** Raw Pan-UKBB summary statistics were downloaded into disease-specific subdirectories (e.g., phenocode-153-colorectal-cancer/).
- **Script:** Custom Unix/Bash extractions (utilizing zgrep and awk), i.e., extract_ukbb_hcar_locus_ver2.sh.
- **Action:** Slices the massive whole-genome files down to the exact topological boundaries of the *HCAR* tandem regulatory array (Chromosome 12: 122.85 Mb – 123.51 Mb; hg19). Dynamically reformats headers and calculates standard *P*-values from the UKBB -log_10_(*P*) formats.
- **Outputs:** Lightweight target files (e.g., UKBB_HCAR_Locus_Extracted_phecode-153-both_sexes_chr12.tsv).

**Phase 3: Cross-Trait Harmonization & Pleiotropy Alignment**

- **Script:** cancer_vs_scz_cross_reference.py
- **Inputs:**
  - Tandem_HCAR2_HCAR1_wide_regulatory.tsv (The baseline SCZ risk architecture—and its genomic extraction was described elsewhere, doi: 10.64898/2026.06.26.734782).
  - The newly extracted Pan-UKBB oncology locus files.
- **Process:** Merges the datasets based on exact genomic coordinates (hg19). Algorithmically aligns alleles to account for reference/alternate strand flips between the two biobanks. Calculates pleiotropic directionality by comparing the directional effect sizes (Betas) of the SCZ risk alleles against the cancer risk alleles.
- **Output:** Cross_Ref_UKBB_HCAR_Locus_Extracted_[disease].tsv (The final analytical matrices used for quadrant plotting, isolating shared vs. protective variants).

**Phase 4: Visualization & Modeling**

This folder contains the Python scripts used to generate the manuscript's primary figures. Outputs are saved to the plots/ directory.

- figure_1_oncology_miami_plotter.py: Generates mirrored locus-zoom plots (Miami Plots) contrasting the massive structural variance of the Vanguard (SCZ) genome against the strict "Statistical Desert" observed in the solid tumor cohorts.
- figure_2_antagonistic_quadrant_fixed.py: Maps the Beta-Beta alignment of the cross-referenced datasets. Visually segregates variants by their topological domains (3' *HCAR1* vs. 5' *HCAR2*) to prove the lactate-specific protective "seesaw" effect.
- figure_3_oncology_schematic.py: Generates the base SVG architectural model detailing the biophysical mechanism of macrophage "lactate blindness" within the tumor microenvironment. (Figure_3_Oncology_Schematic.svg)

**Usage and Reproducibility**

All scripts require standard Python 3 scientific libraries (pandas, numpy, matplotlib, seaborn).

*Note on Raw Data:* Due to file size constraints, the raw, whole-genome .bgz files from the Pan-UKBB are excluded from this repository. They can be re-acquired at any time using the links generated in Phase 1. The lightweight, sliced locus files are provided herein to allow for immediate reproducibility of the cross-trait algorithms and figure generation.
