## Supplementary figures and images for "Antagonistic Pleiotropy at the HCAR1 Locus Reveals a Genetic Basis for Solid Tumor Resistance in Schizophrenia"

### Figure_1_Oncology_vs_SCZ_Miami_Plot.png

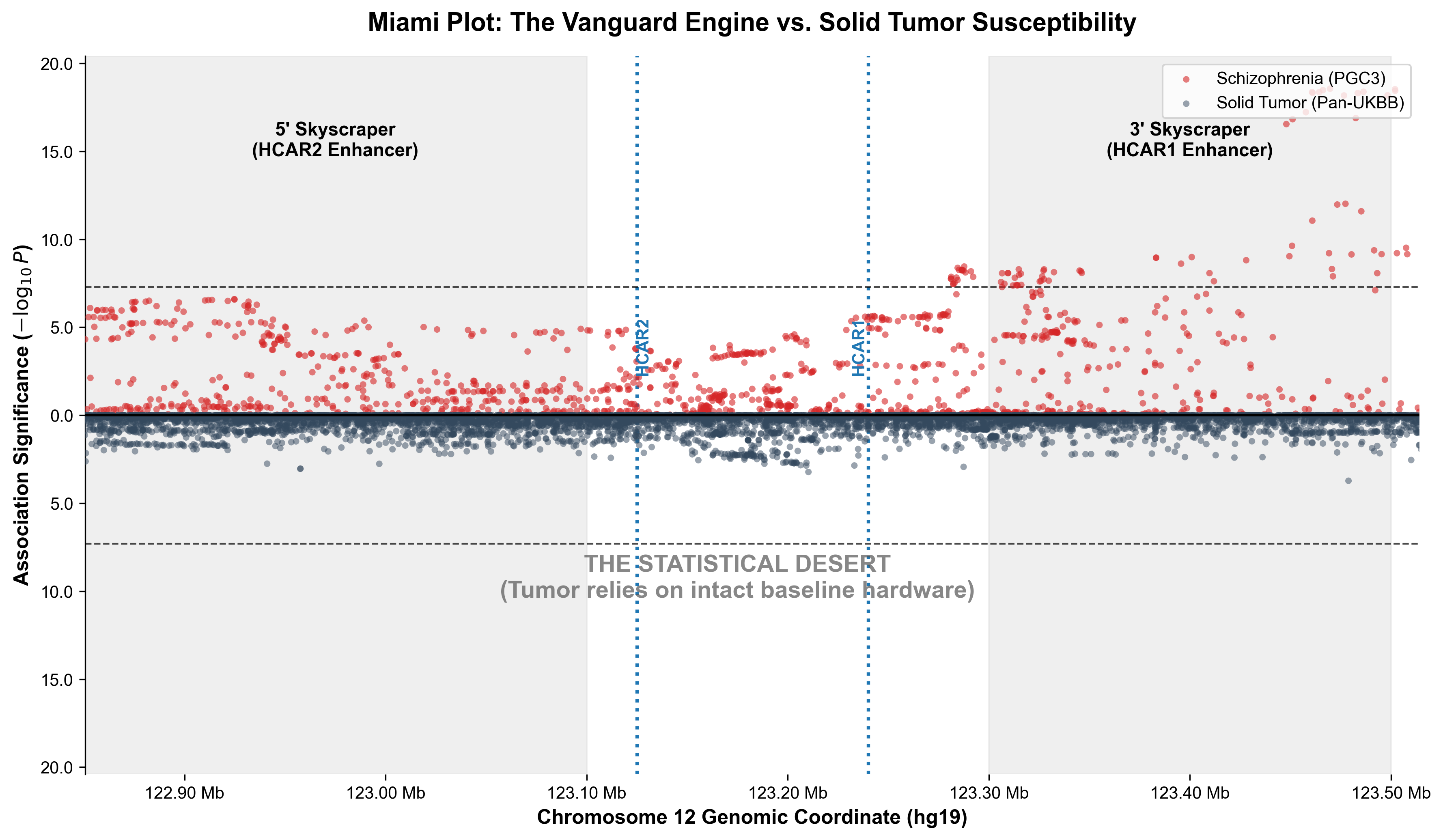

### Figure_2_Antagonistic_Quadrant_Fixed.png

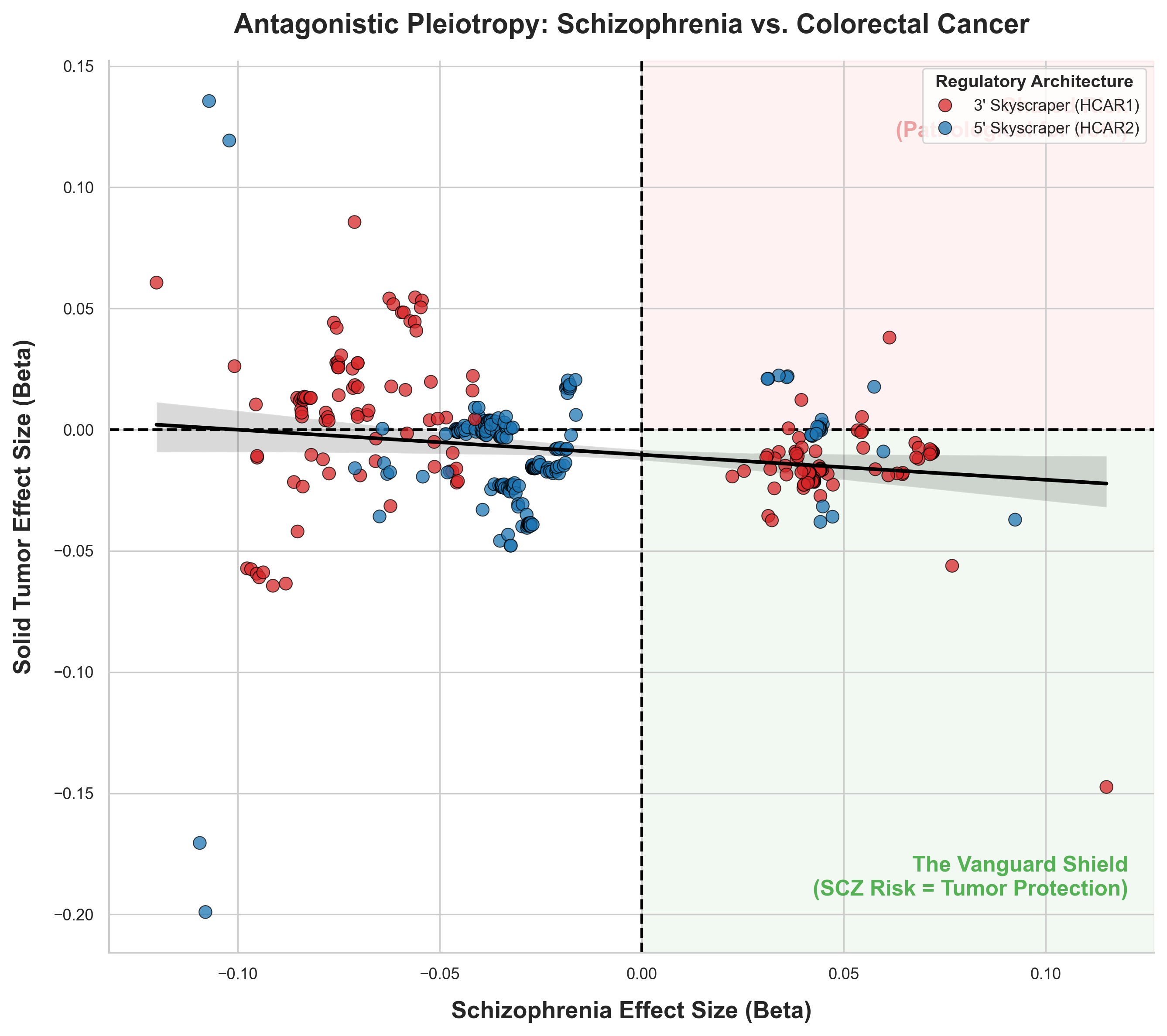
